# Humoral Antibody Kinetics with ChAdOx1-nCOV (Covishield^TM^) and BBV-152 (Covaxin^TM^) Vaccine among Indian Healthcare workers: A 6-month Longitudinal Cross-sectional Coronavirus Vaccine-induced Antibody Titre (COVAT) Study

**DOI:** 10.1101/2022.02.03.22270182

**Authors:** Awadhesh Kumar Singh, Sanjeev Ratnakar Phatak, Ritu Singh, Kingshuk Bhattacharjee, Nagendra Kumar Singh, Arvind Gupta, Arvind Sharma

**Author notes:** Corresponding Author: A. K. Singh, G.D Hospital & Diabetes Institute, Kolkata - 700013, India; e mail. **Declarations of interest:** We wish to confirm that there are no known conflicts of interest associated with this publication and there has been no financial support for this work. **Authorship:** All authors meet the International Committee of Medical Journal Editors (ICMJE) criteria for authorship and take responsibility for the integrity of the work. They confirm that this paper will not be published elsewhere in the same form, in English or in any other language, including electronically.

## Abstract

**Background and Aims:** There is limited data available on longitudinal humoral antibody dynamics following two doses of ChAdOx1-nCOV (Covishield^TM^) and BBV-152 (Covaxin^TM^) vaccine against SARS-CoV-2 among Indians.

**Methods:** We conducted a 6-month longitudinal study in vaccinated healthcare workers by serially measuring quantitative anti-spike antibody at 3-weeks, 3-months and 6- months after the completion of second dose. Geometric mean titer (GMT) and linear mixed models were used to assess the dynamics of antibody levels at 6 months.

**Results:** Of the 481 participants, GMT of anti-spike antibody decreased by 56% at 6- months regardless of demographics and comorbidities in 360 SARS-CoV-2 naive individuals, significantly in hypertensives. Participants with past infection had significantly higher GMT at all time points compared to naive individuals. Among SARS-CoV-2 naive cohorts, a significantly higher GMT was noted amongst the Covishield recipients at all time points, but there was a 44% decline in GMT at 6- month compared to peak titer period. Decline in GMT was insignificant (8%) in Covaxin recipients at 6-month despite a lower GMT at all time points vs. Covishield. There was 5.6-fold decrease in seropositivity rate at 6-month with both vaccines. Participants with type 2 diabetes mellitus have a lower seropositivity rate at all the time points. While seropositivity rate was significantly higher with Covishield vs. Covaxin at all time points except at 6-month where Covaxin recipients had a higher seropositivity, although no difference in seropositivity was noted in propensity-matched analysis.

**Conclusions:** There is waning humoral antibody response following two doses of either vaccine at six months.

**Highlights:**

- We assessed humoral antibody dynamics following two doses of the two vaccines used in India until 6 months.
- Our study of 481 health care workers showed a significant decrease in the anti-spike antibody at 6-months.
- Reduction in antibody was regardless of demographics, comorbidities and the vaccine type.
- T2DM cohorts had lowest seropositivity, while hypertensive had significant antibody decline at 6-month.

## 1. Introduction

Vaccination against Severe Acute Respiratory Syndrome Coronavirus 2 (SARS-CoV-2) infection causing Coronavirus disease 2019 (COVID-19) with ChAdOx1- nCOV (Covishield^TM^) and BBV-152 (Covaxin^TM^) in India started from January 16, 2021 following Emergency Use Approval (EUA) by the Drug Controller General of India. ChAdOx1-nCOV or AZD1222 or Covishield^TM^, acquired from Oxford University and AstraZeneca, manufactured by Serum Institute of India, Pune, is a recombinant replication-deficient chimpanzee adenovirus-vectored vaccine encoding SARS-CoV-2 spike antigen produced in genetically modified human embryonic kidney (HEK) 293 cells. BBV-152 or Covaxin^TM^ manufactured by Bharat Biotech, Hyderabad in collaboration with Indian Council of Medical Research, India, is a ß-propiolactone inactivated whole virion vaccine having all SARS-CoV-2 proteins adjuvanted with imidazoquinoline, a Toll-like receptor 7/8 (TLR 7/8) agonist, to boost the immune response. While each dose (0.5 ml) of Covishield contains 5 x 10^10^ viral spike particles, each 0.5 ml dose of Covaxin contains 6 µg dose of whole virion inactivated corona virus protein of strain NIV- 2020-770. The exact proportion of spike antigen in Covaxin is not exactly known. Available phase 3 randomized clinical trials (RCTs) of both vaccines found them safe and significantly effective [1, 2]. However, there is still a paucity of data in the real-world settings as to how much and how long both these novel vaccines can elicit an immune response both at humoral and cellular level. Long-term antibody kinetics after the completion of both doses of Covishield and Covaxin in Indians is even less well known. We have recently reported the short-term anti-spike antibody humoral response after the first and second dose of both vaccines from Cross-sectional Coronavirus Vaccine-induced Antibody Titre (COVAT) study [3]. Here, we report a longitudinal 6-month follow-up of humoral antibody kinetics from COVAT study after the completion of the second dose.

## 2. Methods

### 2.1 Study design and participants

Our report follows the Strengthening the Reporting of Observational Studies in Epidemiology (STROBE) reporting guideline for cross-sectional studies [4]. COVAT study was a pan-India, cross-sectional study approved by the ethical committee of Thakershy Charitable Trust, Ahmedabad, Gujarat, India. Informed consent was taken on Google-sheet from all the participants who volunteered to participate in this study. Inclusion and exclusion criteria for this study has already been published earlier [3]. Summarily, all adult health care workers (HCWs) of >18 years of age who completed two dose of either vaccine and had completed a total of four measured anti-spike antibody titre until 6-month of second dose were included in this analysis. Measurement of anti-spike antibody at four time-points include – a. first sample: day 21 after the first dose until the day before the second dose, b. second sample: day 21-28 of second dose, c. third sample: day 83-97 (3- months) of second-dose and, d. fourth sample: day 173-187 (6-months) after the second dose. An additional 7 days for collection of blood samples was allowed due to sporadic lockdown at that point of time. Participants who had past COVID-19 (recovered from the COVID-19 >6 weeks before the first dose of either vaccine) were also included in this study. Participants who acquired confirmed SARS-CoV- 2 infection <6-week before the first dose of vaccine, and between first dose and within 2-weeks of second dose of vaccine were excluded from this analysis.

### 2.2 Measurements

All samples were collected as either serum or plasma using EDTA vials from each participant and analyzed at Central laboratory of Neuberg, Supratech at Ahmedabad, Gujarat, India. The IgG antibodies to SARS-CoV-2 directed against the spike protein (S-antigen, both S1 and S2 protein) were assayed with LIASON^®^ S1/S2 quantitative antibody detection kit (DiaSorin Saluggia, Italy) using indirect chemiluminescence immunoassay (CLIA). The diagnostic sensitivity of this kit has been reported to be 97.4% with a specificity of 98.5% as per manufacturer’s protocol. Antibody levels >15.0 arbitrary unit (AU)/mL were considered as sero-positive, while antibody level ≤15 AU/mL were considered as seronegative, as per the manufacturer’s kit. This kit has found to have a concordance to neutralizing antibody (NAb) done by Plaque Reduction Neutralization Test (PRNT) with a positive agreement of 94.4% and negative agreement of 97.8% at a cut-off of >15 AU/mL. The lower and upper limit of this quantitative spike antibody kit is 3.8 and 400 AU/mL respectively, as per the manufacture’s brochure [5].

Clinical data was collected from all eligible participants including age, sex, blood groups, body mass index (BMI), past history of confirmed SARS-CoV-2 infection, any comorbidities, presence of diabetes mellitus (type 1 [T1DM] and type 2 [T2DM]), hypertension (HTN) including its duration and treatment received, dyslipidemia, ischemic heart disease (IHD), chronic kidney disease (CKD) and cancer. Data was collected for any adverse events post-vaccination and subsequent SARS-CoV-2 infection including breakthrough infections.

### 2.3 Statistical analysis

Standard descriptive statistics were used to present the demographic characteristics of patients included in this study. Categorical data are shown in counts and percentages whereas quantitative antibody levels data are presented in geometric mean titer (GMT) with 95% confidence interval (CI). Differences in demographic and clinical characteristics between groups were computed using Chi-square test for categorical variables. Linear mixed models were fitted to examine the anti-spike antibody kinetics over the 6-month period after receipt of the second vaccine dose and to associate these changes with the demographic characteristics and co- existing conditions of the study participants. The dependent variable was first log-transformed. The covariates in the linear mixed model analysis included sex, age group (< 60 years and > 60 years), body-mass index (BMI; the weight in kilograms divided by the square of the height in meters), co-existing co-morbid conditions, diabetes status, duration of diabetes, hypertension status, duration of hypertension, dyslipidemia, ischemic heart disease, type of vaccine etc. Time was modeled as a linear trend from the receipt of the second dose up to 6 months. The estimated effects of covariates are presented as ratios of means with 95% confidence intervals on the original logarithmic scale of the anti-spike antibodies. Cochran’s Q test is used to determine if there are differences on a dichotomous dependent variable i.e., the seropositivity rate (defined as yes or no) between different time points of antibody titer measurement after second dose of vaccination. Repeated measures ANOVA with post-hoc adjustment method is used to evaluate for any significant differences in antibody titer measurements between different time points within similar cohort. We took into account both Greenhouse Geiser correction and Huynh-Feldt correction, in case the assumption of sphericity was violated by Mauchly’s test of sphericity. One-way ANOVA (analysis of variance) with post-hoc test has been used to find the significance of study parameters between three cohort of patients viz. SARS-CoV-2 naïve, breakthrough infection and past infection. A propensity score was generated taking into consideration age, sex and BMI of the SARS-CoV-2 naive cohorts after two complete doses of either vaccine to eliminate the bias that could have creeped in due to the convenience sampling method. Multiple binary logistic regression analysis was carried out to identify the independent predictors of breakthrough infection. A two-sided p value of less than 0.05 was considered to indicate statistical significance. Entire statistical analysis was carried out with Statistical Package for Social Sciences (SPSS Complex Samples) Software Version 22.0 for windows, SPSS Inc., Chicago, IL, USA, with Microsoft Word and Excel being used to generate graphs and tables.

## 3. Results

Data collection for this analysis started since the January 16, 2021 (first day of vaccination amongst HCW) until November 15, 2021 (data-lock date). Of the 515 vaccinated health care workers who were eligible for the study, 481 were selected for this analysis who had completed results from all 4 blood samples until 6-month follow-up, following the completion of second dose. Of the 481 participants, 64 had breakthrough SARS-CoV-2 infection (positive RT-PCR/RAT result for SARS-CoV-2, ≥2-weeks after the second dose), while 57 participant had a confirmed past ≥ history of COVID-19 (6-weeks before the first dose of vaccine). **Supplementary figure 1** summarizes the patients’ disposition for this 6-month longitudinal follow-up analysis. The demographic characteristics and data on coexisting conditions in the study participants are provided in **supplementary table 1** for all the three groups- a. SARS-CoV-2 naive until data-lock date, b. Participants with breakthrough infection and, c. Participant with past history of COVID-19 before first dose of vaccine.

### 3.1 Mixed-model analysis of variables associated with anti-spike antibody titers after two doses of vaccine in SARS-CoV-2 naive cohorts

As mentioned in methodology, a linear mixed model was fitted to examine the anti-spike antibody kinetics over the 6-month period (time was modeled as a linear trend) in SARS-CoV-2 naive individuals (N = 360) to study the change with the demographic characteristics and with associated co-morbidities of the study participants. Our analysis found that in the peak periods (defined as day 21 through 28 after the receipt of second dose of vaccine) a significantly lower anti-spike antibody titers was associated with participants with age >60 years and participants having any co-morbidities compared to age ≤60 years and individuals without comorbidities, but no such difference was noted at the end-of-study period (defined as day 180 through 187 after receipt of the second dose). There was no difference noted in the anti-spike antibody titers at the peak and at the end-of-study period with regard to gender, body-mass index and blood group. While no significant difference in antibody titre was noted in participants with T2DM, dyslipidemia, ischemic heart disease vs. without these comorbidities, a significantly lower anti-spike antibody titer was seen in the hypertensive subjects at the end of study period (but not in the peak period). Notably, while a higher anti-spike antibody titer was noted amongst the Covishield recipients at the peak titer period (factor 1.62; 95% CI, 1.15-2.26), this was no longer significant at the end-of-study period (factor 1.37; 95% CI, 0.77-1.59). Similar findings were noted in the mixed model analysis of propensity matched (age, Sex and BMI) cohorts (n = 41, in each group) who took either Covishield or Covaxin. Propensity matched analysis found a significantly higher anti-spike antibody titre amongst the Covishield recipients during the peak period (factor 1.72; 95% CI, 1.18-2.63) compared to Covaxin but not at the end-of-study period (factor 1.46; 95% CI, 0.83-1.67). **Table 1** summarizes the findings from mixed model analysis.

**Table 1:**
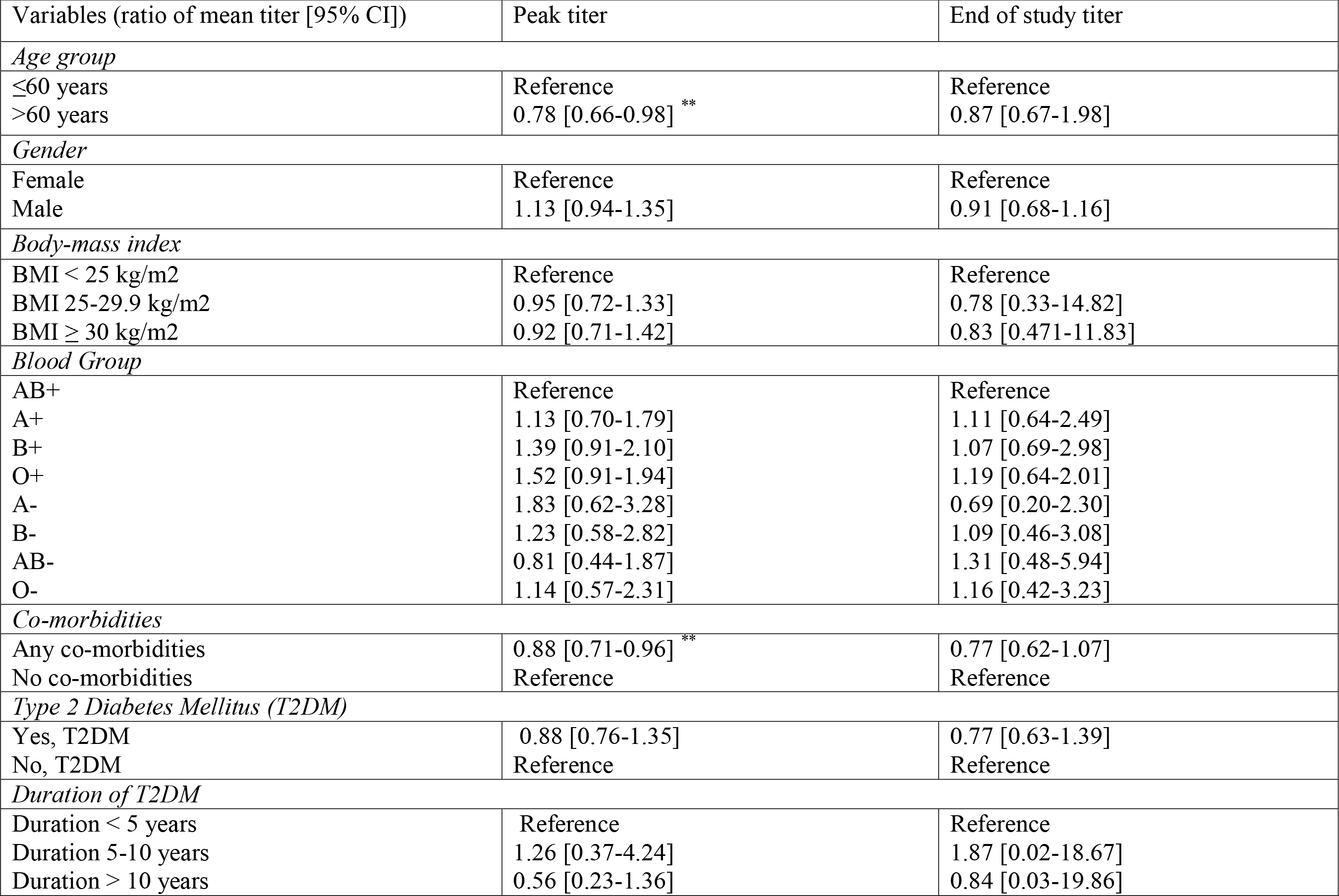
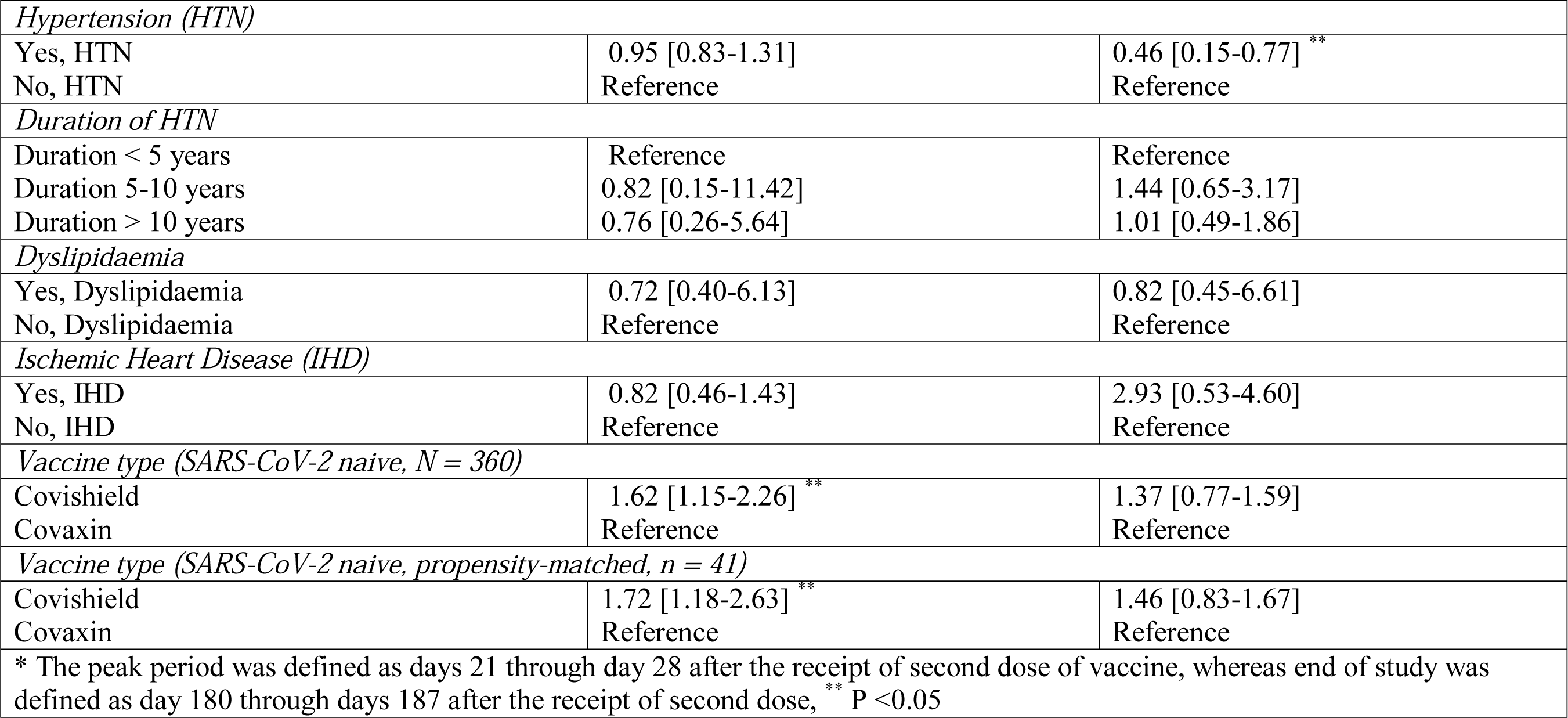
Mixed-model analysis of variables associated with anti-spike antibody titers after following two doses of vaccine in SARS-CoV-2 naïve cohorts

Collectively, there was a trend of decline in antibody titre in all participants over time but it was significantly lower at the end of 6-months in hypertensive vs. normotensive participants.

### 3.2 Trend in anti-spike antibody GMT between SARS-CoV-2 naïve, those with breakthrough infection and individuals with past history of COVID-19 after the first dose of vaccine until 6-months after the second dose

A significant difference (p <0.001) in the geometric mean antibody titre was noted between the participants who were SARS-CoV-2 naive, had breakthrough infection and with a past history of COVID-19 during all the sampling time points as computed by one-way ANOVA test. Amongst all fully vaccinated cohorts, there was a significantly lower GMT in SARS-CoV-2 naive participants at 3- and 6- month (GMT 67.83 AU/mL; 95% CI, 67.24-68.15 AU/mL at 3-month, and GMT 44.77 AU/mL; 95% CI, 44.09-45.23 AU/mL at 6-month) versus those who had past history of COVID-19 (GMT 277.86 AU/mL; 95% CI, 276.19-278.22 AU/mL at 3-month, and GMT 205.24 AU/mL; 95% CI, 204.87-206.31 AU/mL at 6-month; p<0.001 for both time points) or who had breakthrough infection (GMT 338.84 AU/mL; 95% CI, 337.77-339.16 AU/mL at 3-month, and GMT 245.47 AU/mL; 95% CI, 245.29-245.54 AU/mL at 6-month; p<0.001 at both time points). Notably, there was no difference in GMT between patients with past history of COVID-19 and breakthrough infection both at 3-month (p = 0.397) and 6-month (p = 0.369) after the second dose. When we analysed the trend in antibody titre for within-group difference across various time points by repeated measures ANOVA, we found a significant increase in GMT in SARS-CoV-2 naive cohort at 21-days and 3-months after the second dose (both p <0.001) but the GMT returned to the baseline value (titre at 21-days after the first dose) at 6-months (p = 0.177). In the breakthrough infection cohort, the GMT increased significantly across all the subsequent sampling (21-days and 3-months post-second dose, both p <0.001), however there was a significant (p <0.001) dip in the antibody titre at 6-months after second dose. For the participants with past history of COVID-19, there was a nominally significant increase in GMT at 21-days after second dose (p = 0.049) compared to 21-days after first dose (baseline value), and no significant difference noted compared to baseline value at 3-months (p = 0.26) and 6-months (p = 0.15) after the second dose. Overall, there was 41% decline in anti-spike GMT at 6- month compared to peak period (21-days after the second dose) in all 481 participants. In SARS-CoV-2 naive individuals, anti-spike GMT decreased by 56% at 6-month compared to the peak period. **Figure 1** depicts the mean GMT at all time points across all groups. For the Covishield recipients, the anti-spike GMT increased significantly from baseline (21-days post-first dose) to 21-days post-second dose (p <0.001) but it dipped significantly at 3- and 6-months post-second dose (p <0.001 for all timepoint comparison). In case of Covaxin recipients, the GMT of antibody increased significantly from baseline (21-days post-first dose) to 21-days post-second dose (p <0.001) but the GMT of antibody plateaued at 3- and 6-months post-second dose (all p >0.05) unlike the Covishield recipient. Comparatively, anti-spike antibody GMT was significantly higher at all time points in Covishield recipients compared to Covaxin recipients, including at 6-months following second dose. Interestingly, while there was a significant 44% decline in the anti-spike antibody GMT in Covishield recipients at 6-months compared to the peak period (21-days after the second dose), no significant (8%) decline in anti-spike GMT was observed in Covaxin recipients at 6-months. This finding is unique in light of significant and consistent greater generation of anti-spike antibody GMT with Covishield compared to the Covaxin at all time points (21-days post-first dose until 6 month post-second dose of vaccine). **Figure 2** depicts the GMT with either vaccine in overall and propensity-matched SARS-CoV-2 naive individuals. **Table 2** summarizes the findings between the three groups.

**Figure 1:**
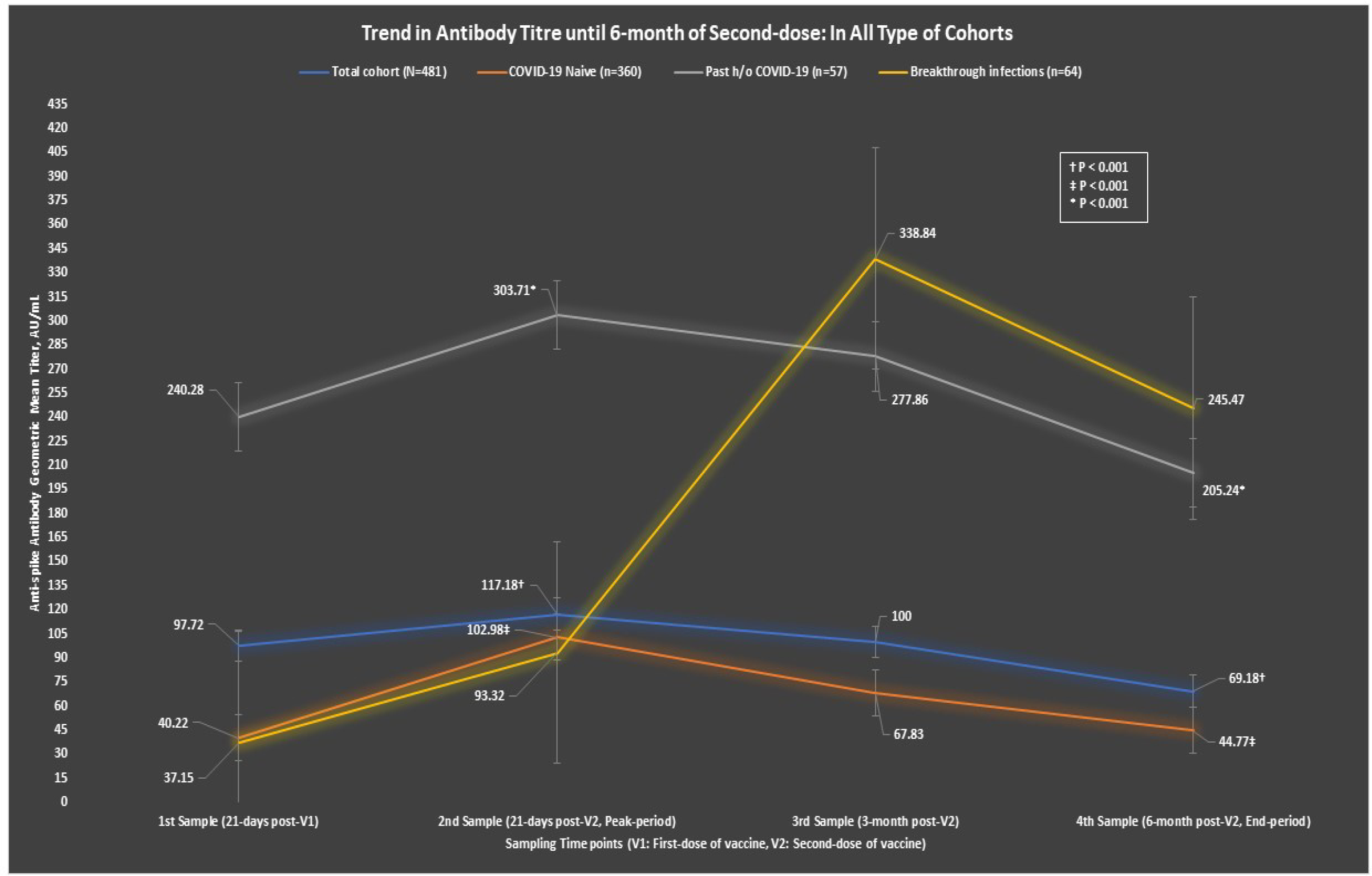
GMT of anti-spike antibody over 6-months in all types (naive vs. convalescent vs. breakthrough infection) of cohort

**Figure 2:**
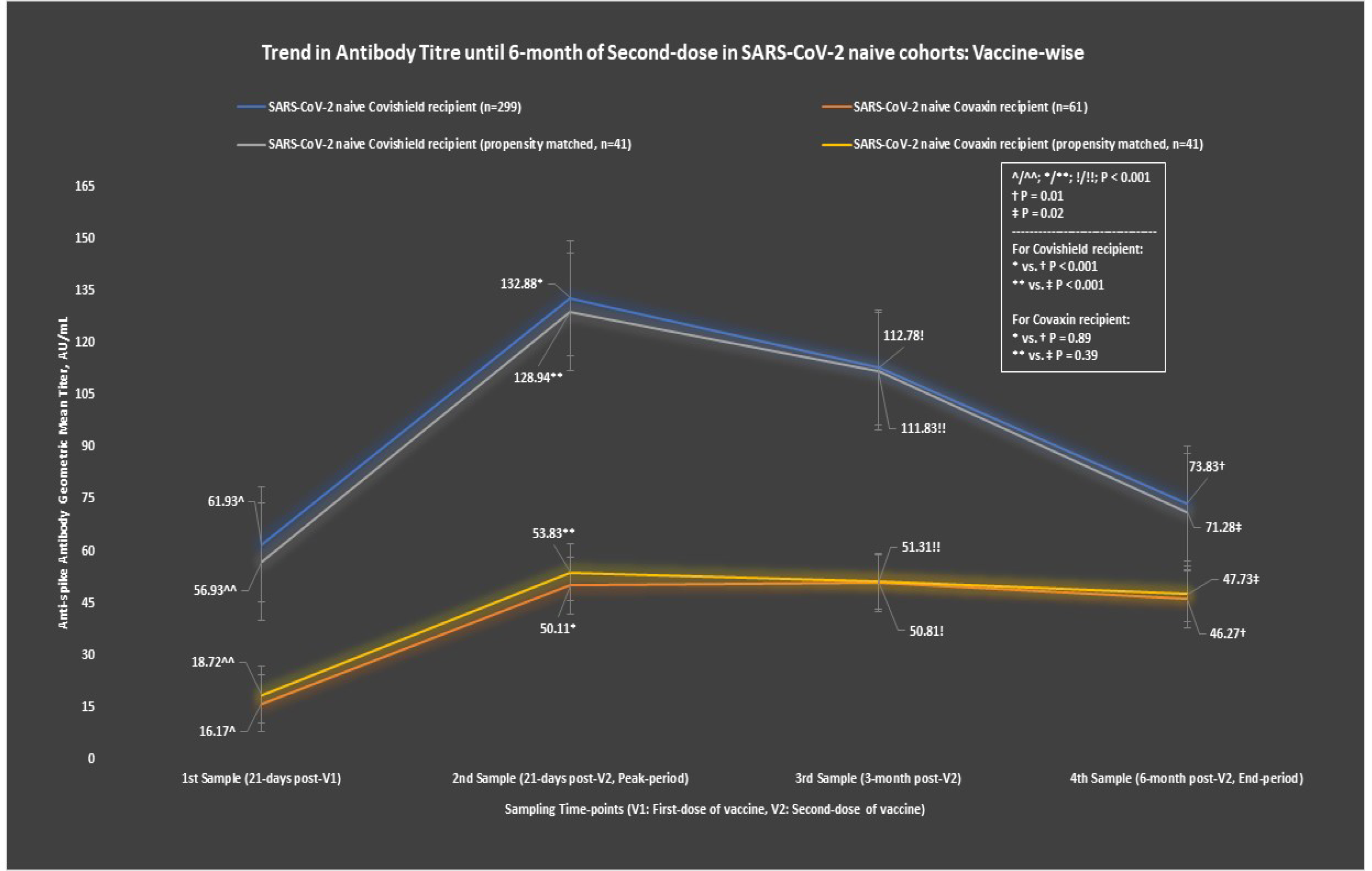
GMT of anti-spike antibody over 6-month with Covishield and Covaxin in all SARS-CoV-2 naive and propensity-matched cohorts

**Table 2:** Geometric mean titre (GMT) of anti-spike antibody in SARS-CoV-2 naive, breakthrough infection and individuals with past history of COVID-19 after first dose of vaccine until 6-month after the second dose:

|  | Antibody titer, Geometric Mean (95% CI) in AU/mL |  |  |  |  |
| --- | --- | --- | --- | --- | --- |
|  | First sample,<br>21 days post-V1 | Second sample,<br>21 days post-V2 | Third sample,<br>3-months post-V2 | Fourth sample,<br>6-months post-V2 | P** |
| Total cohort,<br>N = 481 | 97.72<br>(96.45-97.87) | 117.18<br>(116.54-117.65) | 100<br>(98.94-100.52) | 69.18<br>(68.95-69.33) | P <0.001; d - <0.001, e - <0.001,<br>f - <0.001, g - 0.005, h - <0.001,<br>i - <0.001 |
| SARS-CoV-2 naive,<br>N = 360 | 40.22<br>(39.62-40.89) | 102.98<br>(101.59-103.22) | 67.83<br>(67.24-68.15) | 44.77<br>(44.09-45.23) | P <0.001; d - <0.001, e - <0.001,<br>f - 0.177, g - <0.001, h - <0.001,<br>i - <0.001 |
| Breakthrough<br>infection; N = 64 | 37.15<br>(37.03, 37.26) | 93.32<br>(93.21-93.46) | 338.84<br>(337.77-339.16) | 245.47<br>(245.29-245.54) | P <0.001; d - <0.001, e - <0.001,<br>f - <0.001, g - <0.001, h -<br><0.001, i - <0.001 |
| Past h/o COVID-19;<br>N = 57 | 240.28<br>(238.76-240.01) | 303.71<br>(302.94-304.02) | 277.86<br>(276.19-278.22) | 205.24<br>(204.87-206.31) | P <0.001; d - 0.049, e - 0.262, f -<br>0.146, g - 0.149, h - <0.001,<br>i - <0.001 |
| p* | p <0.001; a - <0.001,<br>b - 0.370, c - <0.001 | p <0.001; a - <0.001,<br>b - 0.684, c - <0.001 | p <0.001; a - <0.001,<br>b - <0.001; c - 0.397 | p <0.001; a - <0.001,<br>b - <0.001; c - 0.369 |  |
| *SARS-CoV-2 naive cohorts, N = 360* |  |  |  |  |  |
| Covishield recipients | 61.93<br>(54.85-69.52) | 132.88<br>(122.88-144.40) | 112.78<br>(100.23-126.89) | 73.83<br>(64.54-84.47) | P <0.001; d - <0.001, e - <0.001,<br>f - 0.012, g - <0.001, h - <0.001,<br>i - <0.001 |
| Covaxin recipients | 16.17<br>(10.96-23.87) | 50.11<br>(37.30-67.31) | 50.81<br>(33.46-77.15) | 46.27<br>(31.18-68.68) | P <0.001; d - <0.001, e - <0.001,<br>f - 0.002, g - 0.201, h - 0.891,<br>i - 0.634 |
| p* | <0.001 | <0.001 | <0.001 | 0.010 |  |
| *Propensity-matched SARS-CoV-2 naive cohorts, N = 41* |  |  |  |  |  |
| Covishield recipients | 56.93<br>(53.74-60.28) | 128.94<br>(124.48-130.65) | 111.83<br>(108.98-124.83) | 71.28<br>(63.4-83.47) | P <0.001; d - <0.001, e - <0.001,<br>f - 0.019, g - <0.001, h - <0.001,<br>i - <0.001 |
| Covaxin recipients | 18.72<br>(14.68-25.11) | 53.83<br>(39.07-66.18) | 51.31<br>(34.96-76.74) | 47.73<br>(34.78-69.92) | P <0.001; d - <0.001, e - <0.001,<br>f - 0.004, g - 0.840, h - 0.392, i-0.296 |
| p* | <0.001 | <0.001 | <0.001 | 0.021 |  |

p\* < 0.05 considered as statistically significant when p computed by ANOVA test followed by post-hoc Bonferroni test; a: p value between SARS-CoV-2 naive and past h/o COVID-19, b: p value between SARS-CoV-2 naive and breakthrough infection, c: p value between past h/o COVID-19 and breakthrough infection; AND,
P\*\* < 0.05 considered as statistically significant when p computed by repeated measures ANOVA followed by post-hoc test; d: p value between first and second samples, e: p value between first and third samples, f: p value between first and fourth samples, g: p value between second and third samples, h: p value between second and fourth samples, i: p value between third and fourth samples.

Collectively, these findings suggest a greater antibody response following vaccination in those who had SARS-CoV-2 infection either before the first dose or after the second dose compared to infection naive individuals, which is an expected finding. Importantly, this longitudinal study also showed that anti-spike antibody decreased by 56% at 6-months as compared to the peak period in SARS-COV-2 naive cohorts. Despite a significantly greater GMT of anti-spike antibody in Covishield recipients at all time point compared to Covaxin in SARS-CoV-2 naive cohorts, there was a significant 44% decline in anti-spike antibody in the former group compared to the later (nonsignificant 8%). And, a similar significant decline (45%) with Covishield while no apparent decline (11%) with Covaxin was also noted in propensity matched (N =41, in each arm) SARS-CoV-2 naive cohorts.

### 3.3 Trend in seropositivity rate in SARS-CoV-2 naive cohorts

Overall, there was a significant (5.6-fold) decline in the seropositivity rate over a period of six months after the second dose in SARS-CoV-2 naive cohort. Reduction in seropositivity at the 6-month was noted in relation to all the studied clinical demographic parameters (p <0.001) regardless of the type of vaccine received. The seropositivity rate in the age group ≤60 years is significantly higher at all the time points compared to the age >60 years. Similarly, participants with T2DM have a lower seropositivity rate at all the time points compared to those without. No difference in seropositivity was noted at any of the time points with regard to gender, BMI, blood group, presence or absence of HTN (including its duration), dyslipidemia and IHD. Although Covishield recipients have higher seropositivity rate both at 21-days and 3-months after the second dose, a significantly (P = 0.009) higher seropositivity rate in Covaxin recipients (38%) was noted at six months compared to Covishield recipient (22%). However, in the propensity-matched analysis there was no significant (p = 0.24) difference in seropositivity rate at 6-months between the two vaccines. **Table 3** summarizes the findings on trend of seropositivity rate in SARS-CoV-2 naive individuals.

**Table 3:** Trend in seropositivity rate in SARS-CoV-2 naive cohort

| Variable | Second sample,<br>21 days post-V2 | Third sample,<br>3-months post-V2 | Fourth sample,<br>6-months post-V2 | P* |
| --- | --- | --- | --- | --- |
| *Overall seropositivity, n/N (%)* | 342/360 (95.0) | 311/360 (86.4) | 259/360 (71.9) | <0.001 |
| *Age* |  |  |  |  |
| ≤ 60 years, n/N (%) | 295/306 (96.4) | 271/306 (88.6) | 238/306 (77.8) | <0.001 |
| > 60 years, n/N (%) | 47/54 (87) | 40/54 (74.1) | 21/54 (38.9) | <0.001 |
| P<sup>#</sup> value | 0.010 | 0.008 | 0.016 |  |
| *Sex* |  |  |  |  |
| Female, n/N (%) | 142/148 (95.9) | 130/148 (87.8) | 34/148 (23) | <0.001 |
| Male, n/N (%) | 200/212 (94.3) | 181/212 (85.4) | 55/212 (25.9) | <0.001 |
| P<sup>#</sup> value | 0.625 | 0.536 | 0.537 |  |
| *Body-mass index (BMI)* |  |  |  |  |
| BMI < 25 kg/m<sup>2</sup>, n/N (%) | 245/257 (95.3) | 221/257 (86) | 60/257 (23.3) | <0.001 |
| BMI 25-29.9 kg/m<sup>2</sup>, n/N (%) | 59/63 (93.7) | 56/63 (88.9) | 17/63 (27) | <0.001 |
| BMI ≥ 30 kg/m<sup>2</sup>, n/N (%) | 38/40 (95) | 34/40 (85) | 12/40 (30) | <0.001 |
| P<sup>#</sup> value | 0.860 | 0.798 | 0.597 |  |
| *Co-morbidities* |  |  |  |  |
| Any Co-morbidities, n/N (%) | 78/84 (92.9) | 67/84 (79.8) | 29/84 (34.5) | <0.001 |
| No Co-morbidities, n/N (%) | 264/276 (95.7) | 244/276 (88.4) | 60/276 (21.7) | <0.001 |
| P<sup>#</sup> value | 0.389 | 0.048 | 0.021 |  |
| *Blood group* |  |  |  |  |
| A+ve, n/N (%) | 69/74 (93.2) | 67/74 (90.5) | 20/74 (27) | <0.001 |
| B+ve, n/N (%) | 96/100 (96) | 87/100 (87) | 26/100 (26) | <0.001 |
| AB+ve, n/N (%) | 17/19 (89.5) | 15/19 (78.9) | 7/19 (36.8) | <0.001 |
| O+ve, n/N (%) | 137/144 (95.1) | 122/144 (84.7) | 31/144 (21.5) | <0.001 |
| A-ve, n/N (%) | 6/6 (100) | 6/6 (100) | 1/6 (16.7) | <0.001 |
| B-ve, n/N (%) | 8/8 (100) | 6/8 (75) | 3/8 (37.5) | <0.001 |
| AB-ve, n/N (%) | 2/2 (100) | 2/2 (100) | 0/2 (0) | <0.001 |
| O-ve, n/N (%) | 7/7 (100) | 6/7 (85.7) | 1/7 (14.3) | <0.001 |
| P<sup>#</sup> value | 0.389 | 0.197 | 0.080 |  |
| *Type 2 diabetes mellitus (T2DM)* |  |  |  |  |
| Yes, T2DM, n/N (%) | 31/37 (83.8) | 27/37 (73) | 16/37 (43.2) | <0.001 |
| No, T2DM, n/N (%) | 311/323 (96.3) | 284/323 (87.9) | 250/323 (77.4) | <0.001 |
| P<sup>#</sup> value | 0.006 | 0.017 | 0.007 |  |
| *Duration of DM* |  |  |  |  |
| Duration < 5 years, n/N (%) | 6/6 (100) | 5/6 (83.3) | 2/6 (33.3) | <0.001 |
| Duration 5-10 years, n/N (%) | 16/21 (76.2) | 14/21 (66.7) | 10/21 (47.6) | <0.001 |
| Duration > 10 years, n/N (%) | 9/10 (90) | 8/10 (80) | 4/10 (40) | <0.001 |
| P<sup>#</sup> value | 0.001 | 0.046 | 0.042 |  |
| *Hypertension (HTN)* |  |  |  |  |
| Yes, HTN, n/N (%) | 62/67 (92.5) | 52/67 (77.6) | 21/67 (31.3) | <0.001 |
| No, HTN, n/N (%) | 280/293 (95.6) | 259/293 (88.4) | 68/293 (23.2) | <0.001 |
| P<sup>#</sup> value | 0.348 | 0.029 | 0.208 |  |
| *Duration of HTN* |  |  |  |  |
| Duration < 5 years, n/N (%) | 22/22 (100) | 18/22 (81.8) | 5/22 (22.7) | <0.001 |
| Duration 5-10 years, n/N (%) | 28/32 (87.5) | 24/32 (75) | 12/32 (37.5) | <0.001 |
| Duration > 10 years, n/N (%) | 12/13 (92.3) | 10/13 (76.9) | 4/13 (30.8) | <0.001 |
| P<sup>#</sup> value | 0.148 | 0.116 | 0.325 |  |
| *Dyslipidaemia* |  |  |  |  |
| Yes, Dyslipidaemia, n/N (%) | 18/19 (94.7) | 15/19 (78.9) | 6/19 (31.6) | <0.001 |
| No, Dyslipidaemia, n/N (%) | 324/341 (95.0) | 296/341 (86.8) | 83/341 (24.4) | <0.001 |
| P<sup>#</sup> value | 0.973 | 0.579 | 0.662 |  |
| *Ischemic Heart Disease (IHD)* |  |  |  |  |
| Yes, IHD, n/N (%) | 8/8 (100) | 6/8 (75) | 3/8 (37.5) | <0.001 |
| No, IHD, n/N (%) | 334/352 (94.9) | 305/352 (86.6) | 86/352 (24.4) | <0.001 |
| P<sup>#</sup> value | 0.999 | 0.299 | 0.414 |  |
| *Vaccine type* |  |  |  |  |
| Covishield, n/N (%) | 295/299 (98.7) | 277/299 (92.6) | 66/299 (22.1) | <0.001 |
| Covaxin, n/N (%) | 47/61 (77) | 34/61 (55.7) | 23/61 (37.7) | <0.001 |
| P<sup>#</sup> value | <0.001 | <0.001 | 0.009 |  |
| *Vaccine type (Propensity matched)* |  |  |  |  |
| Covishield, n/N (%) | 39/41 (95.12) | 37/41 (90.24) | 11/41 (26.82) | <0.001 |
| Covaxin, n/N (%) | 31/41 (75.61) | 24/41 (58.54) | 16/41 (39.02) | <0.001 |
| P# value | 0.012 | 0.001 | 0.240 |  |
| P* computed by Cochran's Q test;<br>P# computed by chi-square test or Fischer's Exact test;<br>Anti-spike antibody levels >15.0 arbitrary unit (AU)/mL were considered as seropositive while antibody level ≤15 AU/mL were considered as seronegative, as per the manufacturer's kit; V2- Second dose of vaccine |  |  |  |  |

### 3.4 Breakthrough infections and its correlation to demographic characteristics, comorbidities and vaccine type

Overall, in the span of 6-months, 64 participants had breakthrough infections ≥2-weeks after the second dose of vaccine but all were mild to moderate (definition criteria published earlier [3]) in nature, regardless of the vaccine type. There was no significant difference (p = 0.86) in breakthrough infection rate between Covishield (54/407, 13.3%) and Covaxin (10/74, 13.5%) recipients. Multiple logistic regression analysis to identify the independent predictors could not pin point any demographic or clinical factor that may have any significant association to breakthrough infections including the vaccine type. **Supplementary table 2** summarizes the results from multiple logistic regression analysis.

## 4. Discussion

In this cross-sectional longitudinal study, there was a significant waning of humoral response within 6 months after receipt of the second dose of both Covishield and Covaxin vaccine in 360 SARS-CoV-2 naive cohort of HCWs, regardless of age, sex, BMI and comorbidities. While we observed a 56% decrease in anti-spike antibody GMT at 6-month after an initial peak following completion of second dose, hypertensive cohorts had significantly less GMT at all time points vs. normotensive participants. Similarly, people with T2DM had significantly less seropositivity to anti-spike antibody at all time points. Our findings are in concordance to several other studies conducted with different vaccines administered globally. A prospective longitudinal study (N = 3,808) from Israel found a similar rapid decline in humoral response following BNT162b2 (Pfizer-Biotech) vaccine within 6-months after the two complete doses [6]. Another 6- month study of BNT162b2 (Pfizer-Biotech) vaccine from Israel (N = 2,653) demonstrated a decrease in antibody titer by up to 38% each subsequent month, while 16.1% subjects had antibody levels below the seropositivity threshold at 6- month after the two complete doses [7]. Similar waning of antibody was seen in a 20-week sentinel surveillance study (N = 33,533) conducted with inactivated CoronaVac vaccine from Chile [8]. In a cross-sectional longitudinal study of 552 participants from England and Wales there was a significant decline in anti-spike antibody both with ChAdOx1 (five-fold decline) and BNT162b2 (two-fold decline) at 10-weeks after two complete doses of either vaccine [9]. From the Indian vaccine study standpoint, a prospective longitudinal study from India involving 122 HCWs found a substantial decline in levels of anti-spike antibody (S1 subunit) at 6-month compared to the peak values (one month) after the second dose of the Covishield vaccine, suggesting a sharper decay rate of 72% [10]. Another longitudinal cohort study involving 533 HCWs that compared humoral response in both Covishield and Covaxin recipients found a 2- and 4-fold decrease in anti-spike antibody titer respectively, at 6 months, when measured as median value (95% IQR) [11]. Contrarily, few other studies showed different results with other vaccines. A sustained GMT of anti-spike antibody was observed at the 6-month despite some decline when compared to the initial peak after the two doses of mRNA-1273 (Moderna) vaccine in 33 healthy adults [12]. A study of 20 participants who took one or two doses of Ad26.COV2.S (Johnson & Johnson) vaccine showed a durable humoral and cellular responses at 8-months [13]. A comparative 8-month kinetic study showed differential immune responses induced by two mRNA and Ad26.COV2.S (Johnson & Johnson) vaccines. While both

BNT162b2 (Pfizer-Biotech) and mRNA-1273 (Moderna) vaccines had evoked higher peak anti-spike antibody after the two-doses initially (Moderna > Pfizer-Biotech), it declined sharply by 8 months. In contrast, while one dose of Ad26.COV2.S (Johnson & Johnson) vaccine induced lower antibody response initially, it was relatively stable over the 8-months with minimal-to-no decline [14].

Our 6-month longitudinal cross-sectional study found that people with past history of COVID-19 had significantly higher anti-spike antibody GMT at all time points compared to SARS-CoV-2 naive individuals. This finding is concordant to several other studies on the durability of humoral response in convalescent persons and found anti-spike antibody levels decrease only modestly at 8 to 10 months after the infection but remained significantly higher compared to SARS-CoV-2 naive individuals [6, 15–17]. Our study also found only a nominal increase in anti-spike antibody GMT at the peak period (21-days of second dose) in participants with past history of COVID-19, while no significant GMT difference was noted at 3- and 6-months when compared to baseline GMT at 21-days after the first dose. This may suggest that second dose of vaccine did not boost anti-spike antibody GMT further compared to the first dose in participants who had a past history of COVID-19. However, this finding remains speculative in the absence of single dose vaccine comparator arm in our study. Nevertheless, a small study (N = 59) from USA (Rush University, Chicago) that compared one vs. two-dose of BNT162b2 (Pfizer-Biotech) vaccine in people with past history of COVID-19 found a significantly higher anti-spike antibody after one dose compared with two doses in SARS-CoV-2 naive individuals and the second dose did not significantly increase anti-spike antibody compared to the first dose [17]. We also demonstrated that despite a significant increase in anti-spike antibody GMT with Covishield at all time points (21-days post-first dose to 6-month post-second dose) compared to Covaxin, there was a 44% decline in GMT over 6 months from the initial peak (21- days post first dose) in Covishield recipients. However, in Covaxin recipients anti-spike antibody GMT was relatively stable over the 6-months with minimal-to-no (8%) decline despite a modest rise in anti-spike GMT at all time points. Our finding is in contrary to the only other longitudinal study of Covaxin in Indian HCWs that found a 4-fold decrease in median anti-spike antibody titer at 6-month whilst Covishield recipient had only 2-fold decline despite a significantly higher median antibody titer with Covishield at all time points vs. Covaxin [11]. As mentioned earlier, other Indian study reported a 72% decay rate in antibody titre at 6-month in Covishield recipient [10]. The reason for this discrepancy of slow decay of antibody with Covaxin in our study is not exactly known and needs to be studied, however, however at least one comparative Indian study found Covaxin generating better cell-mediated immunity compared to Covishield whilst Covishield was more effective in inducing humoral immunity compared to Covaxin [18].

From an immunological standpoint, an eventual decay of neutralizing antibody titers over the time following vaccination are not unexpected. A decline of neutralizing antibody levels of 5 to 10 % with each passing year was also seen following vaccination against mumps, measles and rubella [19, 20]. Contrarily, a persistent increase in memory B cells, long lived plasmablast and germinal B cell responses over at least six months have been shown after mRNA vaccination [21, 22]. It should be noted that while good humoral response in the form of neutralizing antibody titer may predict some level of protection from symptomatic infection, cellular immunity is equally or perhaps more important in long term protection against severe disease [23]. From the vaccine effectiveness standpoint, wanning of humoral antibody would likely translate to reduction of vaccine efficacy. Indeed, a recent study of two dose of BNT162b2 vaccine showed a decrease in protection by a factor of four in a period of 4-7 months compared to its efficacy within <2-months after the second dose [24, 25]. While waning humoral response at 6-month with these vaccines justify the need of boosters for antibody mediated protection, no conclusive evidence is yet available that suggest boosters augment the memory B and T cell responses for long term protection against severe disease [22, 26].

To the best of our knowledge, this Pan-India cross-sectional COVAT study would be the first of its kind that has involved HCWs from 13 States and 22 cities and reporting a longitudinal anti-spike antibody kinetics 6-month after the second dose of two different vaccines in three different groups of participants- that include individuals who remained SARS-CoV-2 naive, participants with breakthrough infections and people who had past history of COVID-19 before the first dose of vaccine. However, we do acknowledge several limitations. First, we have used a convenience sampling in the present study which amounts to selection bias. Second, our study had all health care workers, mostly healthy persons and therefore they may not represent the general population. Third, we did not have dilution and neat values for those sample that hit both the upper and lower limit of kit value that assessed anti-spike antibody. This could have likely underestimated the GMT in those recipients who had undiluted plateaued value. Fourth, using the mixed model linear analysis may lead to loss in power to detect the significant difference due to double fitting of the study parameter. Fifth, we used a binary logistic regression to identify the independent predictors of breakthrough infection but this model may miss out any predictor variable which may have non-linear relationship with the outcome variable. Lastly, we have measured only anti-spike binding antibody and could not assess NAb and cell-mediated immune response such as Th-1 and Th-2 dependent antibody or cytokines (primarily due to the lack of standardized commercial labs in India). Although a high correlation with spike protein-based ELISA and different antibody classes to NAb have been documented recently [27]. Finally, because of logistic issues (due to lockdown) during pandemic we could not measure the baseline anti-spike antibody titre prior to the first dose of vaccine.

## Conclusions

In conclusion, this cross-sectional 6-month longitudinal study found- a) Waning of anti-spike antibody titre over time regardless of age, sex, BMI, comorbidities and type of vaccine but it was significantly low in hypertensive cohort vs. normotensive at 6-month, b) Participants with past history of COVID-19 had significantly higher anti-spike antibody titer at all time points compared to SARS-CoV-2 naive individuals after two doses of either vaccine, c) Second dose of vaccine did only a little augmentation of spike antibody titre compared to the first dose, d) Covishield recipients had significantly higher anti-spike antibody titer at all time points until 6-month after two doses, e) There was a 56% decline in anti-spike antibody GMT at 6-months compared to the peak period (21-days after second dose) in SARS-CoV-2 naive individuals, f) There was a 44% decline in anti-spike antibody GMT at 6-months compared to peak period (at 21-days) after the second dose of Covishield, g) There was minimal-to-no reduction (8% decline) in anti-spike antibody GMT at 6-months compared to peak period (at 21-days) after the second dose of Covaxin and similar findings were noted in propensity-matched cohorts, h) There was 5.6-fold reduction in seropositivity to anti-spike antibody at 6-month in SARS-CoV-2 naive cohorts regardless of the vaccine received, i) Despite a significant decline in seropositivity with both vaccines, seropositivity was significantly higher with Covaxin compared to Covishield at 6- month in SARS-CoV-2 naive cohorts, although no significant difference was noted in propensity-matched cohorts, j) No significant difference in breakthrough infection rate was noted between Covishield and Covaxin recipients at 6-month.

## Data Availability

All data produced in the present study are available upon reasonable request to the authors.

## Acknowledgments

We would like to thank all the participants who volunteered for this study. We express our sincere gratitude and acknowledgment to our Indian regional coordinators for the smooth conduct of this study that include (in alphabetical order) – Drs. Akash Kumar Singh (Vadodara), Amit Gupta (Noida), Anuj Maheshwari (Lucknow), Arvind Kumar Ojha (Kolkata), Bhavtharini (Erode), B. Harish Kumar (Mysore), J K Sharma (New Delhi), Jayant Panda (Cuttack), Kavyachand Yalamudi (Guntur), Kiran Shah (Vadodara), M Gowri Sankar (Coimbatore), Manohar KN (Bangalore), Meena Chhabra (New Delhi), Pratap Jethwani (Rajkot), M Shunmugavelu (Trichy), Rajiv Kovil (Mumbai), Sunil Gupta (Nagpur), Subhash Kumar (Patna), Somnath (Hyderabad), Urman Dhruv (Ahmedabad). Our heartfelt thanks to Ms. Roma Dave (Dietician) and Dr. Priya Phatak (Ahmedabad) for keeping entire data up-to-date and confidential at every step. Special thanks to Dr. Bhavini Shah, Dr. Sandip Shah, and Dr. Krutarth Shah from Neuberg Supratech Laboratory, Ahmedabad, for generously supporting our cause.

## Contribution of authors

AKS and SRP conceptualized, and designed the study. NKS, AG and AS monitored the study and captured the data at all points of time. AKS, KB, AS and RS conducted the statistical analysis. AKS and RS wrote the first draft. AKS, KB and RS revised the manuscript. All authors gave their intellectual inputs while preparing the manuscript and agreed mutually to submit to this journal.

## Data sharing

All the authors are responsible for the originality of this study. Original data can be shared from first author, if necessary, after a reasonable request.

---

**Supplementary figure 1:**
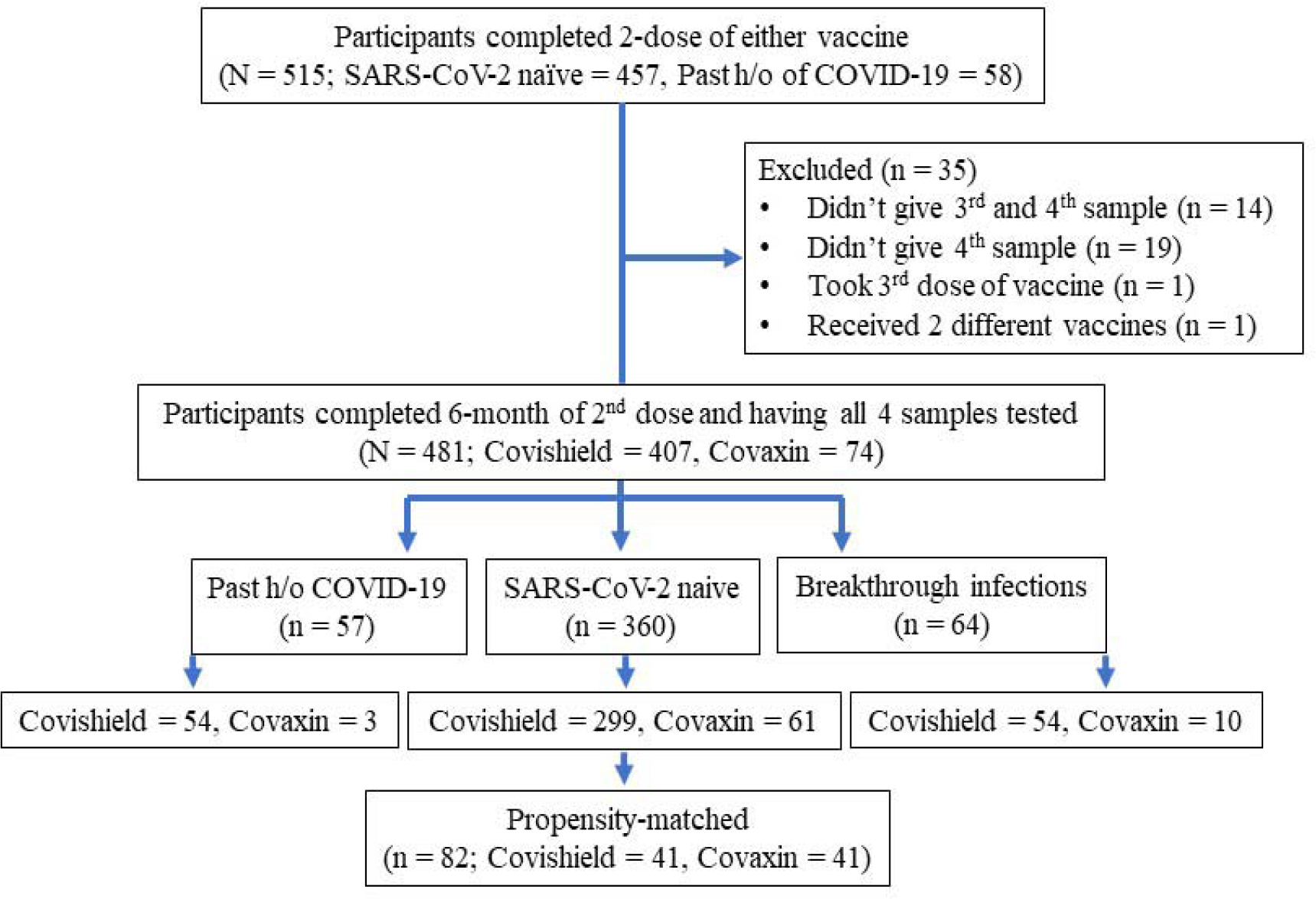
Patients disposition for 6-month longitudinal cross- sectional study

**Supplementary table 1:**
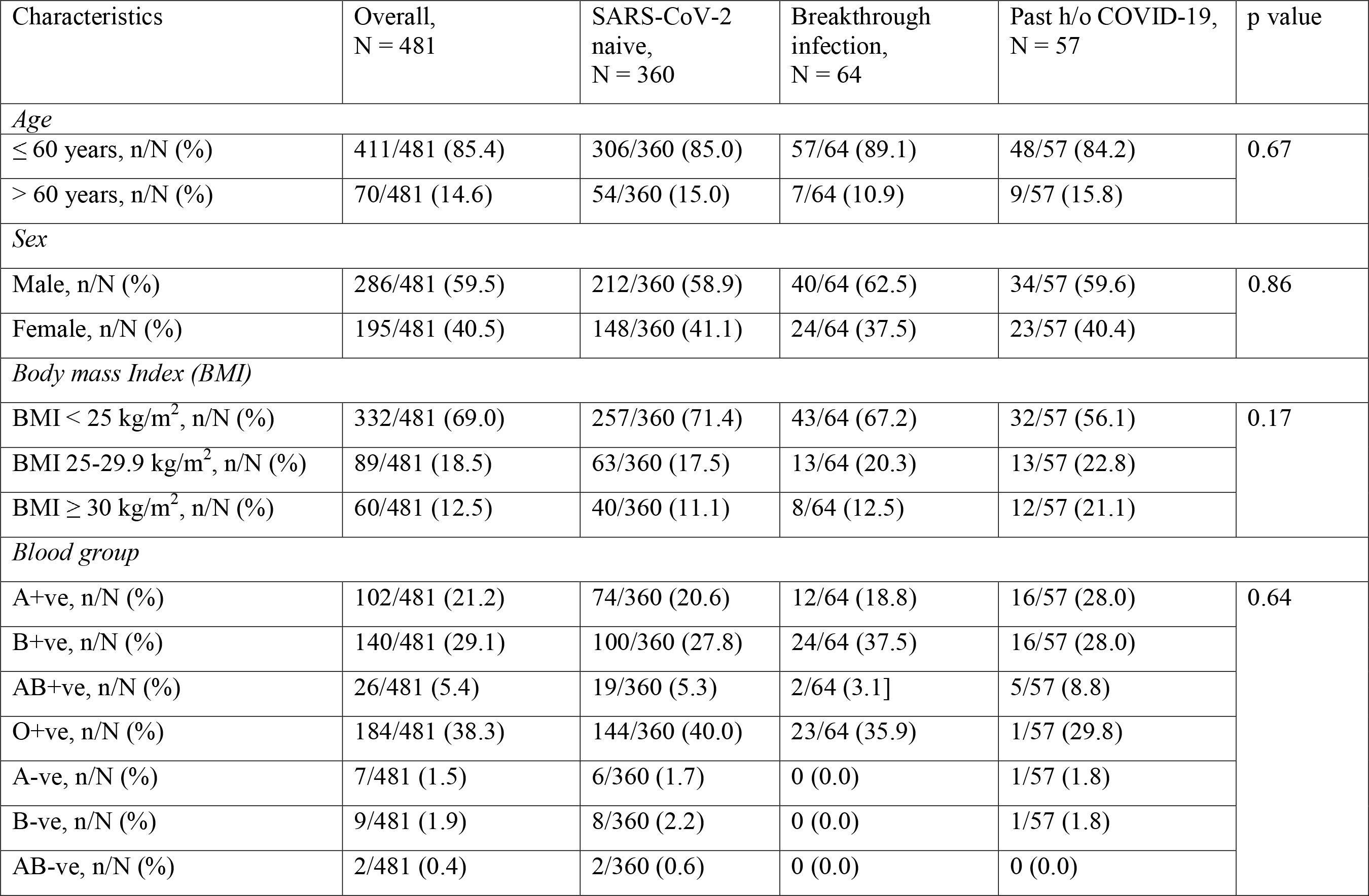
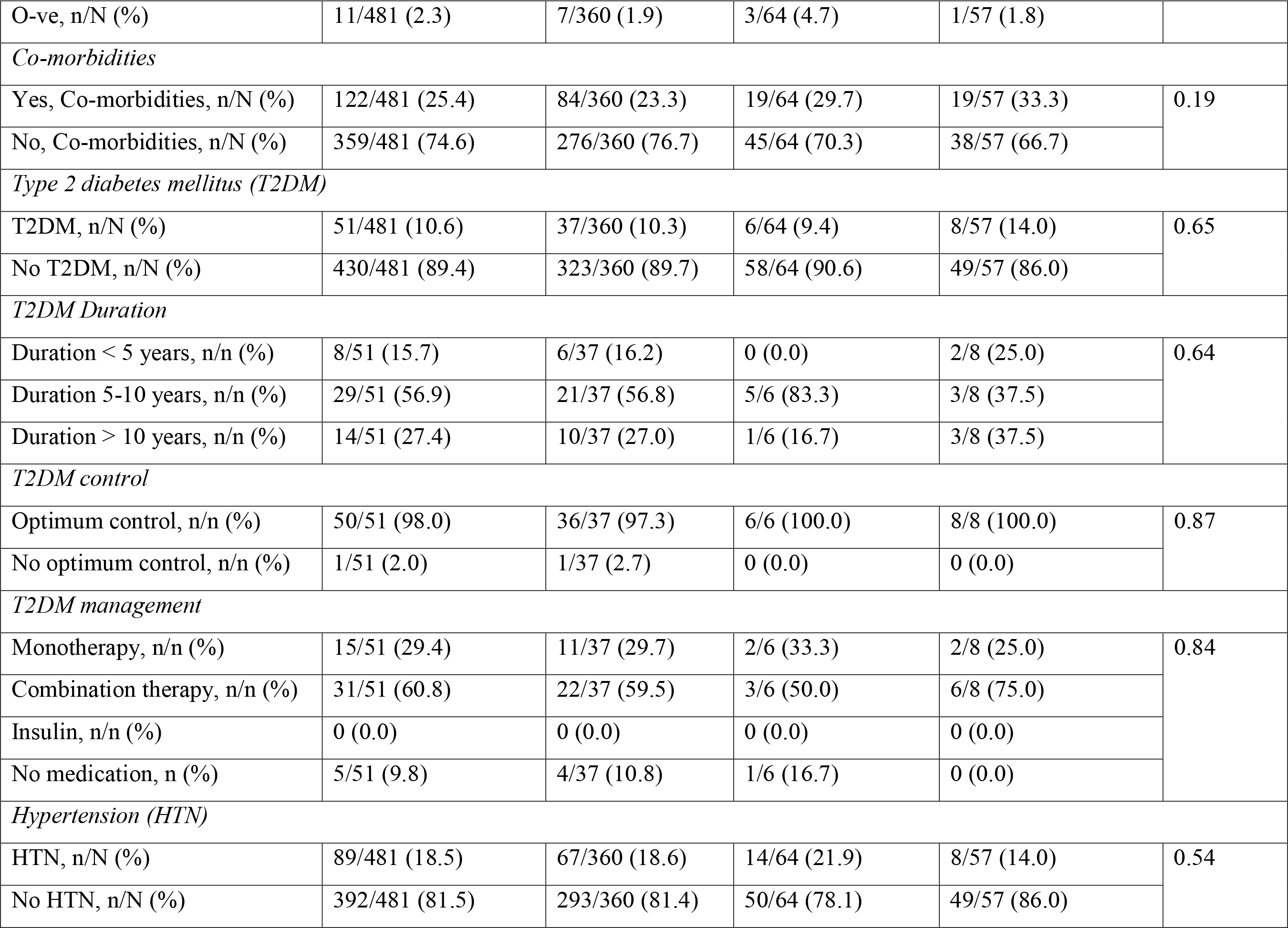
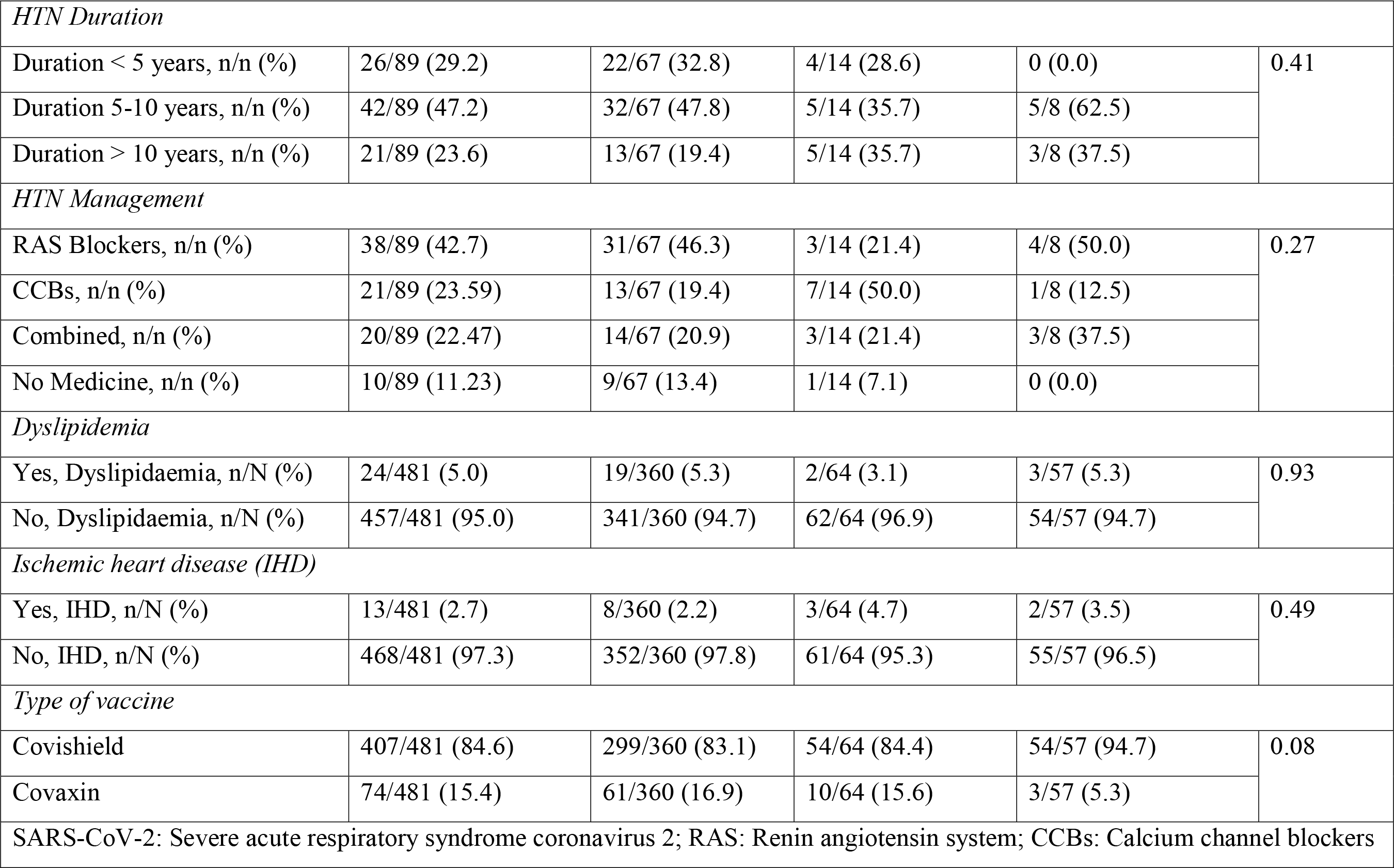
Baseline characteristics of participants in overall and each group

| Characteristics | Overall,<br>N = 481 | SARS-CoV-2<br>naive,<br>N = 360 | Breakthrough<br>infection,<br>N = 64 | Past h/o COVID-19,<br>N = 57 | p value |
| --- | --- | --- | --- | --- | --- |
| *Age* |  |  |  |  |  |
| ≤ 60 years, n/N (%) | 411/481 (85.4) | 306/360 (85.0) | 57/64 (89.1) | 48/57 (84.2) | 0.67 |
| > 60 years, n/N (%) | 70/481 (14.6) | 54/360 (15.0) | 7/64 (10.9) | 9/57 (15.8) |  |
| *Sex* |  |  |  |  |  |
| Male, n/N (%) | 286/481 (59.5) | 212/360 (58.9) | 40/64 (62.5) | 34/57 (59.6) | 0.86 |
| Female, n/N (%) | 195/481 (40.5) | 148/360 (41.1) | 24/64 (37.5) | 23/57 (40.4) |  |
| *Body mass Index (BMI)* |  |  |  |  |  |
| BMI < 25 kg/m<sup>2</sup>, n/N (%) | 332/481 (69.0) | 257/360 (71.4) | 43/64 (67.2) | 32/57 (56.1) | 0.17 |
| BMI 25-29.9 kg/m<sup>2</sup>, n/N (%) | 89/481 (18.5) | 63/360 (17.5) | 13/64 (20.3) | 13/57 (22.8) |  |
| BMI ≥ 30 kg/m<sup>2</sup>, n/N (%) | 60/481 (12.5) | 40/360 (11.1) | 8/64 (12.5) | 12/57 (21.1) |  |
| *Blood group* |  |  |  |  |  |
| A+ve, n/N (%) | 102/481 (21.2) | 74/360 (20.6) | 12/64 (18.8) | 16/57 (28.0) | 0.64 |
| B+ve, n/N (%) | 140/481 (29.1) | 100/360 (27.8) | 24/64 (37.5) | 16/57 (28.0) |  |
| AB+ve, n/N (%) | 26/481 (5.4) | 19/360 (5.3) | 2/64 (3.1] | 5/57 (8.8) |  |
| O+ve, n/N (%) | 184/481 (38.3) | 144/360 (40.0) | 23/64 (35.9) | 1/57 (29.8) |  |
| A-ve, n/N (%) | 7/481 (1.5) | 6/360 (1.7) | 0 (0.0) | 1/57 (1.8) |  |
| B-ve, n/N (%) | 9/481 (1.9) | 8/360 (2.2) | 0 (0.0) | 1/57 (1.8) |  |
| AB-ve, n/N (%) | 2/481 (0.4) | 2/360 (0.6) | 0 (0.0) | 0 (0.0) |  |
| O-ve, n/N (%) | 11/481 (2.3) | 7/360 (1.9) | 3/64 (4.7) | 1/57 (1.8) |  |
| *Co-morbidities* |  |  |  |  |  |
| Yes, Co-morbidities, n/N (%) | 122/481 (25.4) | 84/360 (23.3) | 19/64 (29.7) | 19/57 (33.3) | 0.19 |
| No, Co-morbidities, n/N (%) | 359/481 (74.6) | 276/360 (76.7) | 45/64 (70.3) | 38/57 (66.7) |  |
| *Type 2 diabetes mellitus (T2DM)* |  |  |  |  |  |
| T2DM, n/N (%) | 51/481 (10.6) | 37/360 (10.3) | 6/64 (9.4) | 8/57 (14.0) | 0.65 |
| No T2DM, n/N (%) | 430/481 (89.4) | 323/360 (89.7) | 58/64 (90.6) | 49/57 (86.0) |  |
| *T2DM Duration* |  |  |  |  |  |
| Duration < 5 years, n/n (%) | 8/51 (15.7) | 6/37 (16.2) | 0 (0.0) | 2/8 (25.0) | 0.64 |
| Duration 5-10 years, n/n (%) | 29/51 (56.9) | 21/37 (56.8) | 5/6 (83.3) | 3/8 (37.5) |  |
| Duration > 10 years, n/n (%) | 14/51 (27.4) | 10/37 (27.0) | 1/6 (16.7) | 3/8 (37.5) |  |
| *T2DM control* |  |  |  |  |  |
| Optimum control, n/n (%) | 50/51 (98.0) | 36/37 (97.3) | 6/6 (100.0) | 8/8 (100.0) | 0.87 |
| No optimum control, n/n (%) | 1/51 (2.0) | 1/37 (2.7) | 0 (0.0) | 0 (0.0) |  |
| *T2DM management* |  |  |  |  |  |
| Monotherapy, n/n (%) | 15/51 (29.4) | 11/37 (29.7) | 2/6 (33.3) | 2/8 (25.0) | 0.84 |
| Combination therapy, n/n (%) | 31/51 (60.8) | 22/37 (59.5) | 3/6 (50.0) | 6/8 (75.0) |  |
| Insulin, n/n (%) | 0 (0.0) | 0 (0.0) | 0 (0.0) | 0 (0.0) |  |
| No medication, n (%) | 5/51 (9.8) | 4/37 (10.8) | 1/6 (16.7) | 0 (0.0) |  |
| *Hypertension (HTN)* |  |  |  |  |  |
| HTN, n/N (%) | 89/481 (18.5) | 67/360 (18.6) | 14/64 (21.9) | 8/57 (14.0) | 0.54 |
| No HTN, n/N (%) | 392/481 (81.5) | 293/360 (81.4) | 50/64 (78.1) | 49/57 (86.0) |  |
| *HTN Duration* |  |  |  |  |  |
| Duration < 5 years, n/n (%) | 26/89 (29.2) | 22/67 (32.8) | 4/14 (28.6) | 0 (0.0) | 0.41 |
| Duration 5-10 years, n/n (%) | 42/89 (47.2) | 32/67 (47.8) | 5/14 (35.7) | 5/8 (62.5) |  |
| Duration > 10 years, n/n (%) | 21/89 (23.6) | 13/67 (19.4) | 5/14 (35.7) | 3/8 (37.5) |  |
| *HTN Management* |  |  |  |  |  |
| RAS Blockers, n/n (%) | 38/89 (42.7) | 31/67 (46.3) | 3/14 (21.4) | 4/8 (50.0) | 0.27 |
| CCBs, n/n (%) | 21/89 (23.59) | 13/67 (19.4) | 7/14 (50.0) | 1/8 (12.5) |  |
| Combined, n/n (%) | 20/89 (22.47) | 14/67 (20.9) | 3/14 (21.4) | 3/8 (37.5) |  |
| No Medicine, n/n (%) | 10/89 (11.23) | 9/67 (13.4) | 1/14 (7.1) | 0 (0.0) |  |
| *Dyslipidemia* |  |  |  |  |  |
| Yes, Dyslipidaemia, n/N (%) | 24/481 (5.0) | 19/360 (5.3) | 2/64 (3.1) | 3/57 (5.3) | 0.93 |
| No, Dyslipidaemia, n/N (%) | 457/481 (95.0) | 341/360 (94.7) | 62/64 (96.9) | 54/57 (94.7) |  |
| *Ischemic heart disease (IHD)* |  |  |  |  |  |
| Yes, IHD, n/N (%) | 13/481 (2.7) | 8/360 (2.2) | 3/64 (4.7) | 2/57 (3.5) | 0.49 |
| No, IHD, n/N (%) | 468/481 (97.3) | 352/360 (97.8) | 61/64 (95.3) | 55/57 (96.5) |  |
| *Type of vaccine* |  |  |  |  |  |
| Covishield | 407/481 (84.6) | 299/360 (83.1) | 54/64 (84.4) | 54/57 (94.7) | 0.08 |
| Covaxin | 74/481 (15.4) | 61/360 (16.9) | 10/64 (15.6) | 3/57 (5.3) |  |
| SARS-CoV-2: Severe acute respiratory syndrome coronavirus 2; RAS: Renin angiotensin system; CCBs: Calcium channel blockers |  |  |  |  |  |

**Supplementary table 2:** Multiple logistic regression to identify the independent predictors for breakthrough infection

| Variables in the Equation |  |  |  |  |  |  |  |  |
| --- | --- | --- | --- | --- | --- | --- | --- | --- |
|  | B | S.E. | Wald | df | P | OR | 95% C.I. for OR |  |
|  |  |  |  |  |  |  | Lower | Upper |
| Age | -.371 | .423 | .768 | 1 | .396 | .696 | .301 | 1.582 |
| Sex | -.147 | .277 | .283 | 1 | .595 | .863 | .502 | 1.485 |
| BMI | .043 | .188 | .052 | 1 | .547 | 1.044 | .722 | 1.509 |
| Associated comorbidities | -.252 | .296 | .726 | 1 | .394 | .777 | .435 | 1.388 |
| Type 2 diabetes | -.156 | .457 | .117 | 1 | .732 | .855 | .349 | 2.094 |
| Hypertension | .244 | .328 | .554 | 1 | .456 | 1.277 | .671 | 2.429 |
| Ischemic heart disease | 1.795 | .970 | 3.428 | 1 | .064 | 6.022 | .900 | 40.291 |
| Dyslipidaemia | .001 | .001 | 2.372 | 1 | .124 | 1.000 | 1.000 | 1.000 |
| Covishield vs. Covaxin | .021 | .370 | .003 | 1 | .954 | 1.021 | .495 | 2.110 |
| Constant | 7.929 | 1.884 | 10.943 | 1 | .0001 | 876.85 |  |  |

a. Variable(s) entered on regression step: Age, Sex, BMI (body mass index), Associated Comorbidities, Type 2 diabetes, Hypertension, Ischemic heart disease, Dyslipidaemia, Past history of COVID-19, Covishield, Covaxin, OR: Odds ratio; CI: Confidence interval; S.E: Standard error

